# Serial interval and transmission dynamics during the SARS-CoV-2 Delta variant predominance in South Korea

**DOI:** 10.1101/2021.08.18.21262166

**Authors:** Sukhyun Ryu, Dasom Kim, Jun-Sik Lim, Sheikh Taslim Ali, Benjamin J. Cowling

**Affiliations:** Konyang University College of Medicine, Daejeon, South Korea; Seoul National University, Seoul, South Korea; The University of Hong Kong, Hong Kong, China; Hong Kong Science and Technology Park, Hong Kong

## Abstract

We estimated mean serial interval and superspreading potential for the predominant Delta variant of SARS-CoV-2. Mean serial intervals were similar with 3.7 and 3.5 days during early and latter periods, respectively. Furthermore, the risk of superspreading events was similar with 23% and 25% of cases seeded 80% of all transmissions.

---

South Korea is in the middle of a fourth community epidemic of severe acute respiratory syndrome coronavirus 2 (SARS-CoV-2) transmission, which is now predominated by the B.1.617.2 lineage (Delta variant) (1, 2). The epidemic size largely depends on epidemiological characteristics such as serial interval distribution and transmissibility (3, 4). However, for the Delta variant of SARS-CoV-2, empirical evidence produced using country-level data is limited. Here, we estimated serial interval distribution, reproductive numbers, and superspreading potential of the SARS-CoV-2 during the Delta variant predominance.

## The study

We obtained line-list data on COVID-19 cases reported by Korean local public health authorities between 11 July 2021 and 1 September 2021. Because the detection rate of the Delta variant accounted for more than 50% of local cases after 25 July 2021 and avoiding right censoring bias, we divided the study period into two periods (Period-1: 11 July 2021–24 July 2021 and Period-2: 25 July 2021–15 August 2021). Overall, 82,671 local cases were obtained during the whole study period, and 19,635 and 34,569 cases were identified in Period-1 and Period-2, respectively. The data included information on contact tracing with other reported cases of COVID-19 (i.e. the case number of infector or infectee) and the dates of symptom onset. The serial interval represents the time between symptom onset for both the infector and the infectee in a transmission chain (3). Based on the line-list information, we reconstructed the transmission pairs by identifying the infector and infectee. We identified 3,728 transmission pairs (1,344 pairs in Period-1 and 2,384 pairs in Period-2) having the date of symptom onset for both infector and infectee. The overall mean and standard deviation (SD) of the serial interval estimate were 3.6 days (95% Credible Interval [CrI]: 3.5, 3.6) and 4.9 days (95% CrI: 4.9, 5.0), respectively. The mean and standard deviation of the serial interval estimate during Period-1 and Period-2 were 3.7 days (95% CrI: 3.5, 3.8) with an SD of 4.8 days (95% CrI: 4.8, 4.9), and 3.5 days (95% CrI: 3.4, 3.6) with an SD of 5.0 days (95% CrI: 4.9, 5.0), respectively (Figure 1A). We used Welch’s two-sample t-test to compare the mean serial interval for Periods-1 and Period-2 and found no significant difference (p-value=0.40).

**Figure 1.**
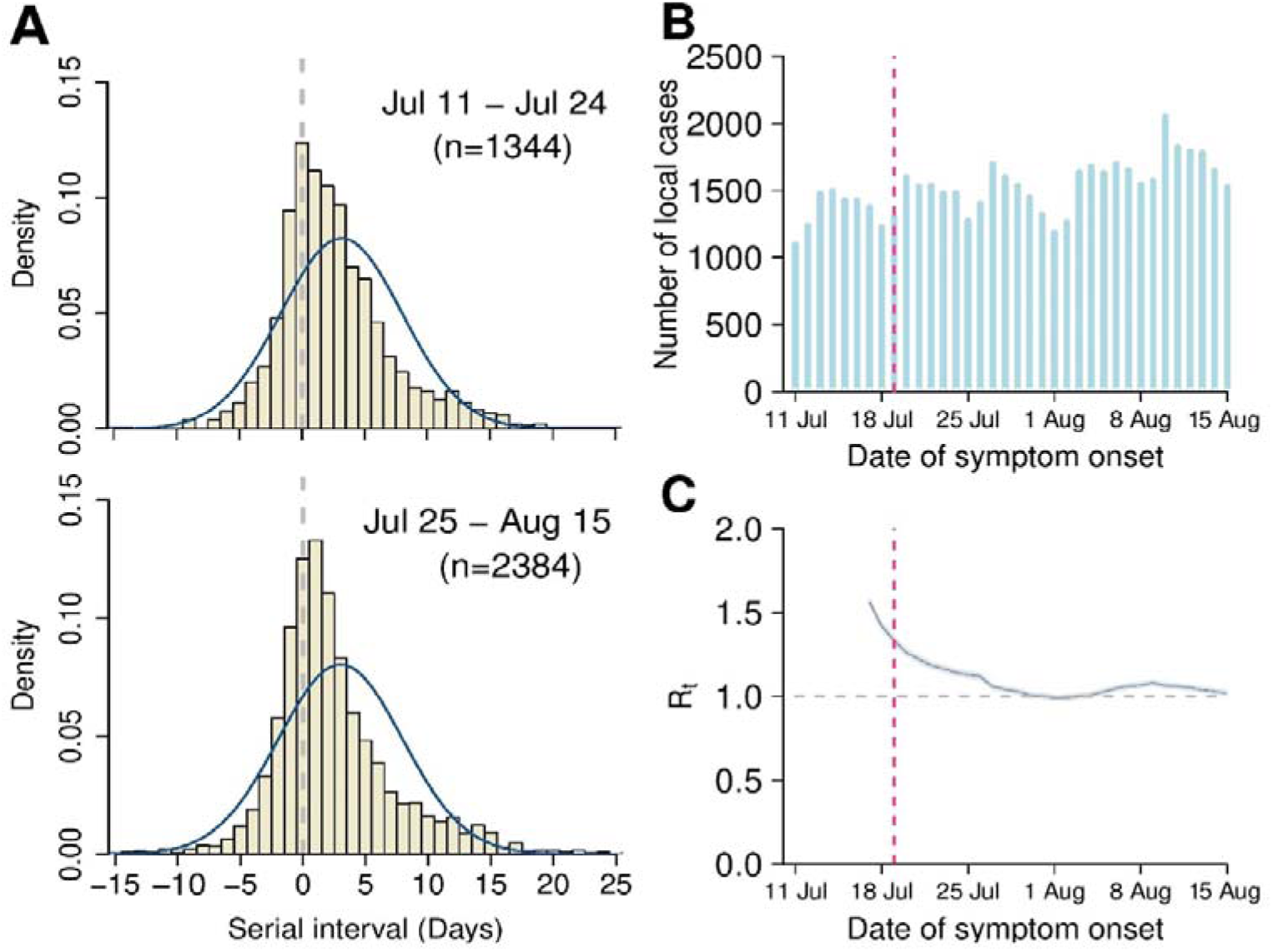
Estimated serial interval distribution, incidence of COVID-19, and transmissibility during the Delta variant of SARS-CoV-2 predominance in South Korea. A) The estimated serial interval distribution by using the 3728 infector-infectee pairs. The vertical bars indicate the distribution of empirical serial intervals, and the solid blue line indicates the fitted normal distribution. B) Reported number of confirmed COVID-19 cases by the date of symptom onset. The red vertical dashed line indicates the date of implementation of an enhanced social distancing including limiting gathering sizes to 4 people nationwide (19 July 2021). C) Estimated daily *R*_*t*_ of SARS-CoV-2 (blue line) with 95 credible intervals (gray shade). The gray horizontal dashed line indicates the critical threshold of *R*_*t*_ =1. The red vertical dashed line indicates the date of implementation of an enhanced social distancing. *R*_*t*_ =effective reproductive number

To identify the potential changes in SARS-CoV-2 transmissibility, we estimated the time-varying effective reproductive number (*R*_*t*_), which defines the mean number of secondary infectious cases generated from a typical primary infectious case at time *t*. The epidemic becomes under control if *R*_*t*_ falls below 1 sustainably. We estimated *R*_*t*_ using the *EpiEstim* package in R (5). In South Korea, nonpharmaceutical interventions including the nationwide mask mandate have been implemented since 2020. As a large number of COVID-19 cases were identified on the mid of July 2021, limiting gathering sizes to 4 people was implemented from 19 July 2021 nationwide (6) (Figure 1B and 1C). However, we identified that the estimated *R*_*t*_ was sustained above 1 during the study period (Figure 1C).

For the analysis of superspreading potential, we identified 5,778 transmission pairs included non-symptom onset for either infector or infectee (2,169 pairs for Period-1 and 3,609 pairs for Period-2). We calculated the number of secondary cases for each individual from the transmission pairs and fitted the data into a negative binomial distribution (7) (Appendix). The two parameters of the distribution represent the reproduction number (*R*) and overdispersion parameter (*k*), respectively. The estimated *k* of Period-1 and Period-2 was 0.64 (95% CrI: 0.57, 0.72) and 0.85 (95% CrI: 0.75, 0.98), which corresponded to an expected proportion of cases responsible for 80% of secondary cases of 23% (95% CrI: 22%, 24%) and 25% (95% CrI: 24%, 26%) (Figure 2).

**Figure 2.**
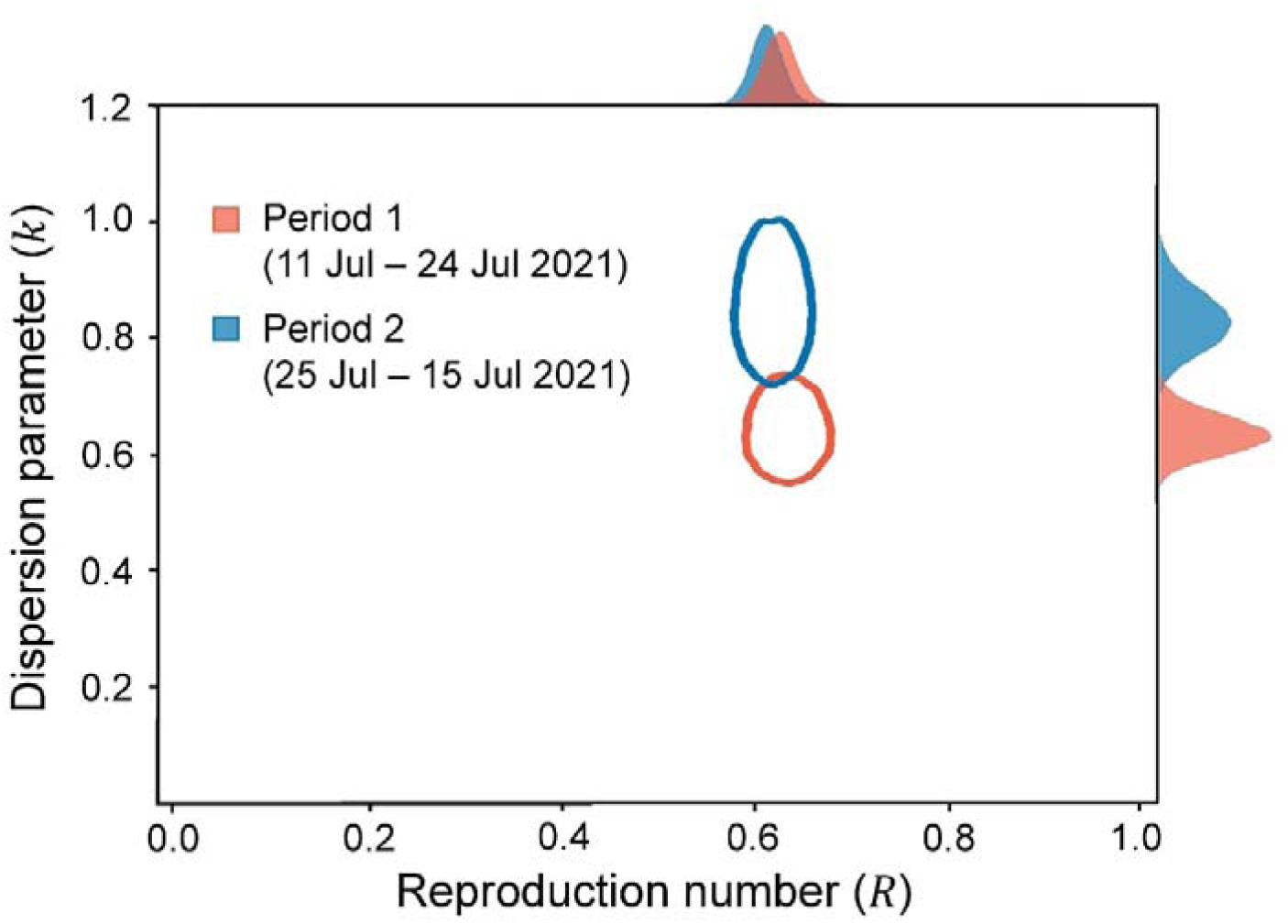
Risk of superspreading events for COVID-19 during the Delta variant of SARS-CoV-2 predominance in South Korea. Joint estimates of the overdispersion parameter (*k*) and reproduction number (*R*) of COVID-19 by using 5,778 pairs (2,169 for Period-1 and 3,609 for Period-2). The red and blue ovals indicate the bivariate 95% credible region of the estimated overdispersion parameter and reproduction number for Period-1 and Period-2. The posterior marginal distributions were plotted in red and blue shaded regions. Note: Period-1, 11 July 2021 – 24 July 2021; Period-2, 25 July 2021 – 15 August 2021.

## Conclusions

We estimated the serial interval distributions of SARS-CoV-2 for early and latter periods of the Delta variant predominance, and identified the mean serial intervals were similar across two different periods. This is consistent with a recent study suggesting no significant differences in the serial intervals between patients infected with the Delta variant and the wild type (8). However, in contrast, our finding suggested that the mean serial interval was 1 day longer than the estimates reported in a study describing the faster spread of the Delta variant in China (mean serial interval of 2.3 days) compared to the wild type (9). The changes in the implementation of public health measures such as active contact tracing and rapid isolation of COVID-19 patients would have shortened the serial interval, and reduced transmissibility and superspreading potential (3, 4). However, the South Korean public health authority has consistently implemented strategies for active case finding and the immediate isolation of laboratory-confirmed COVID-19 patients and exposed individuals using digital QR codes since 10 June 2020 (10). Therefore, the impact of enhanced case isolation against the serial interval of SARS-CoV-2 is likely limited in our study. Furthermore, restricting large gatherings had likely reduced the superspreading potential. However, as the *R*_*t*_ was above 1 during most of the study period, the nonpharmaceutical interventions implemented were likely insufficient to control the transmission of SARS-CoV-2 in South Korea.

Our study has some limitations. First, in our analysis, we did not consider the impact of COVID-19 vaccinations. About 14% of transmission pairs used in this study were linked with older adults (≥ 60 years of age), who might have received COVID-19 vaccinations. However, the vaccination program was not implemented to the public below 55 years of age by early August 2021. Second, we did not consider changes in nonpharmaceutical interventions on the local level and the increased amount of travel as the study period covered summer holidays. However, enhanced social distancing including limiting gathering sizes to 4 people was implemented nationwide during the study period. Third, we retrieved online case reports, which could have some inaccuracies in the information. However, the daily number of laboratory-confirmed local cases was similar between the collected line list and official daily reports (Appendix). Lastly, as the individual genotype information was not included in the line-list data, the proportion of the Delta variant was evaluated from alternative data retrieved from the Korea Disease Control and Prevention Agency.

A previous Korean study, which examined the early transmissibility of SARS-CoV-2 in February-March 2020, estimated the mean reproduction number as 1.5 for the wild type (11), and the early epidemic of COVID-19 was successfully controlled with non-lockdown social distancing (12). Our findings suggest that the introduction of the Delta variant is likely to have increased the difficulty of controlling SARS-CoV-2 transmission in South Korea. The large number of COVID-19 cases in South Korea during the study period could be explained by the increased secondary attack rate generated by cases infected with the Delta variant (13, 14), which is in line with a previous study (8). Encouraging COVID-19 vaccination and further strengthening nonpharmaceutical interventions are warranted to mitigate the Delta variant.

## Data Availability

The data is available upon reasonable request to the corresponding author.

## Acknowledgments

This work was supported by the Basic Science Research Program through the National Research Foundation of Korea, funded by the Korean Ministry of Education (NRF-2020R1I1A3066471).

