## Appendix for "Serial interval and transmission dynamics during the SARS-CoV-2 Delta variant predominance in South Korea"

**Contents:**

1. Data
2. Estimating overdispersion parameter ($K$)
3. Supplementary analysis for the risk of superspreading events
4. Appendix Tables
5. Appendix Figures

### 1. Data

We acquired data on COVID-19 cases, confirmed by real-time reverse transcriptase-polymerase chain reaction (RT-PCR) and reported by Korean local public health authorities. The COVID-19 cases included were between 11 July 2021 and 1 September 2021, when the Delta variant accounted for more than 40% of local cases reported from the Korea Disease Control and Prevention Agency (1, 2). The data included contact tracing with other reported cases of COVID-19 and demographic characteristics of the patients, including age, sex, and date of symptom onset (https://github.com/gentryu/COVID-19delta).

### 2. Estimating overdispersion parameter ($K$)

Statistical methods utilized in this study followed our previous COVID-19 study for superspreading events (SSE) (3). The probability function of negative binomial distribution that an index case $(i)$ generate the number of secondary case $(y_i)$ is given by

$$Pr(Y = y_i) = \frac{\Gamma(k + y_i)}{y_i!\,\Gamma(k)}\left(\frac{k}{k + R}\right)^k \left(\frac{R}{k + R}\right)^{y_i},$$

where $k$ and $R$ are the estimated dispersion parameter and reproduction number, respectively, of index case $i$.

To understand the individual variation of infectiousness of COVID-19 during study period, observed offspring distributions were fitted to the negative binomial distribution.

Given the estimated reproduction number and dispersion parameter, the proportion of infected persons responsible for 80% of secondary cases, $P_{80\%}$, was calculated using equations from the previous studies (3-6). The proportion $P_{80\%}$ is given by

$$1 - P_{80\%} = \int_0^X NB\left(\lfloor x \rfloor; k, \frac{k}{R + k}\right) dx,$$

where $X$ satisfies

$$1 - 0.80 = \frac{1}{R}\int_0^X \lfloor x \rfloor \, NB\left(\lfloor x \rfloor; k, \frac{k}{R + k}\right) dx.$$

Furthermore, with the threshold for SSE as 6 secondary cases defined in the previous study (4), the proportion of SSE was estimated for each period with the equations below.

$$\int_6^\infty NB\left(\lfloor x \rfloor; k, \frac{k}{R + k}\right) dx = 1 - \int_0^5 NB\left(\lfloor x \rfloor; k, \frac{k}{R + k}\right) dx.$$

Finally, using the branching process, the expected probability that one index case of SARS-CoV-2 infection results in a cluster of size $s$ were estimated with the equations from previous studies (3, 5, 6) given by

$$r_s = \frac{\Gamma(ks + s - 1)}{\Gamma(ks)\Gamma(s + 1)}\left(\frac{R}{k}\right)^{s-1}\left(\frac{k}{k + R}\right)^{ks + s - 1},$$

where the probability of cluster of size $s$ or greater could be estimated as follows:

$$p_s = 1 - \sum_{l=1}^{s-1} r_l.$$

With the condition that $N$ seed cases were introduced into the totally susceptible populations, the estimated probability that at least one cluster of size $s$ or greater occurs is

$$P_{N,s} = 1 - (1 - p_s)^N.$$

Note that $P_{N,s}$ is equal to $p_s$ when exactly one index case was introduced ($N = 1$).

We used a Bayesian Markov Chain Monte Carlo simulation using *rstan* package. Four chains of 40,000 iterations were obtained with 5,000 burn-in. The prior distributions of dispersion parameter and reproduction number were uniform with lower and upper bounds set at 0 and 100. Convergence was checked visually using a trace plot and the Gelman-Rubin-Brooks diagnostic (7). The posterior distribution of the estimates was demonstrated with the median and 95% credible intervals.

### 3. Supplementary analysis for the risk of superspreading events

We estimated the expected proportion of cases responsible for 80% of the total secondary cases to identify the risk of SSE. Furthermore, the probability of SSEs using estimated $R_0$ and $k$ and the probability that one index case results in a cluster of 10 cases or more were estimated.

Based on the source of infection, we estimate the offspring distribution using infectee-infector pairs. We fitted observed distribution of the secondary case into a negative binomial offspring distribution (Appendix Figure 1). The expected proportion of cases responsible for 80% of secondary cases was 22.99% (95% CrI: 21.97%, 24.03%) and 24.93% (95% CrI: 23.90.71%, 25.97%) for Period 1 and 2, respectively. The probability of SSE and the probability that one index case results in a cluster of 10 cases or more were 0.33% (95% CrI: 0.25%, 0.45%) and 4.71% (95% CrI: 3.92%, 5.60%) for Period-1, and 0.17% (95% CrI: 0.12%, 0.24%) and 4.254% (95% CrI: 3.51%, 5.11%) for Period-2.

### 4. Appendix Tables

**Appendix Table 1.** Age-specific distribution of infector-infectee pairs having symptom onset for both infector and infectee.

| | Age group of infectee, y | | | | | | | | |
|---|---|---|---|---|---|---|---|---|---|
| Age group of infector, y | 0-9 | 10-19 | 20-29 | 30-39 | 40-49 | 50-59 | ≥60 | NA | Total |
| 0-9 | 16 | 16 | 26 | 32 | 20 | 34 | 8 | 1 | 153 |
| 10-19 | 18 | 51 | 82 | 57 | 78 | 45 | 42 | 0 | 373 |
| 20-29 | 48 | 76 | 160 | 119 | 130 | 126 | 52 | 3 | 714 |
| 30-39 | 41 | 57 | 134 | 133 | 103 | 87 | 34 | 2 | 591 |
| 40-49 | 41 | 67 | 112 | 84 | 103 | 115 | 49 | 1 | 572 |
| 50-59 | 34 | 47 | 125 | 96 | 103 | 113 | 60 | 2 | 580 |
| ≥60 | 21 | 31 | 61 | 43 | 43 | 44 | 38 | 1 | 282 |
| NA | 0 | 3 | 2 | 0 | 1 | 5 | 2 | 450 | 463 |
| Total | 219 | 348 | 702 | 564 | 581 | 569 | 285 | 460 | 3728 |

NA: Not available

**Appendix Table 2.** Age-specific distribution of infector-infectee pairs included non-symptom onset for either infector or infectee.

| | Age group of infectee, y | | | | | | | | |
|---|---|---|---|---|---|---|---|---|---|
| Age group of infector, y | 0-9 | 10-19 | 20-29 | 30-39 | 40-49 | 50-59 | ≥60 | NA | Total |
| 0-9 | 36 | 31 | 46 | 58 | 41 | 66 | 19 | 4 | 301 |
| 10-19 | 37 | 62 | 107 | 80 | 109 | 79 | 60 | 0 | 534 |
| 20-29 | 86 | 107 | 234 | 170 | 196 | 179 | 82 | 4 | 1058 |
| 30-39 | 73 | 87 | 195 | 188 | 144 | 136 | 73 | 3 | 899 |
| 40-49 | 64 | 103 | 159 | 127 | 161 | 187 | 75 | 3 | 879 |
| 50-59 | 71 | 84 | 188 | 151 | 139 | 192 | 104 | 3 | 932 |
| ≥60 | 29 | 53 | 117 | 71 | 67 | 79 | 68 | 2 | 486 |
| NA | 0 | 4 | 7 | 0 | 1 | 6 | 4 | 667 | 689 |
| Total | 396 | 531 | 1053 | 845 | 858 | 924 | 485 | 686 | 5778 |

NA: Not available

5. **Appendix Figures**

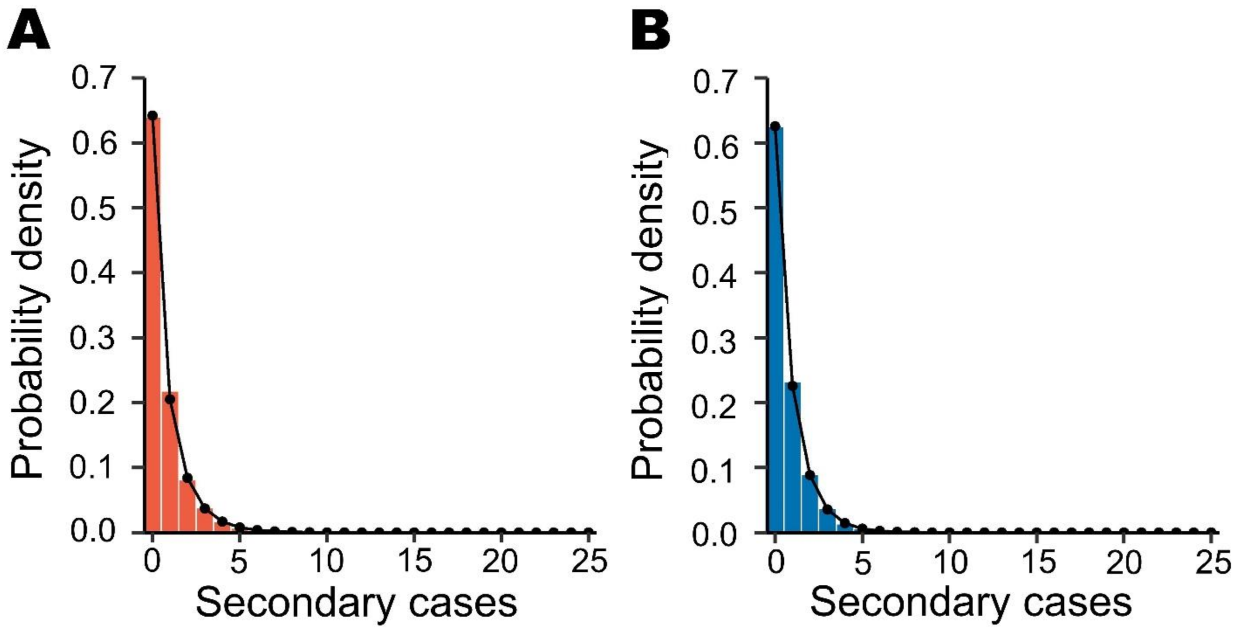


**Appendix Figure 1**. Distributions of the number of secondary cases of COVID-19 in (A) the Period-1 (11 July 2021 – 24 July 2021) and (B) Period-2 (25 July 2021 – 15 August 2021) and corresponding fitted negative binomial distributions.

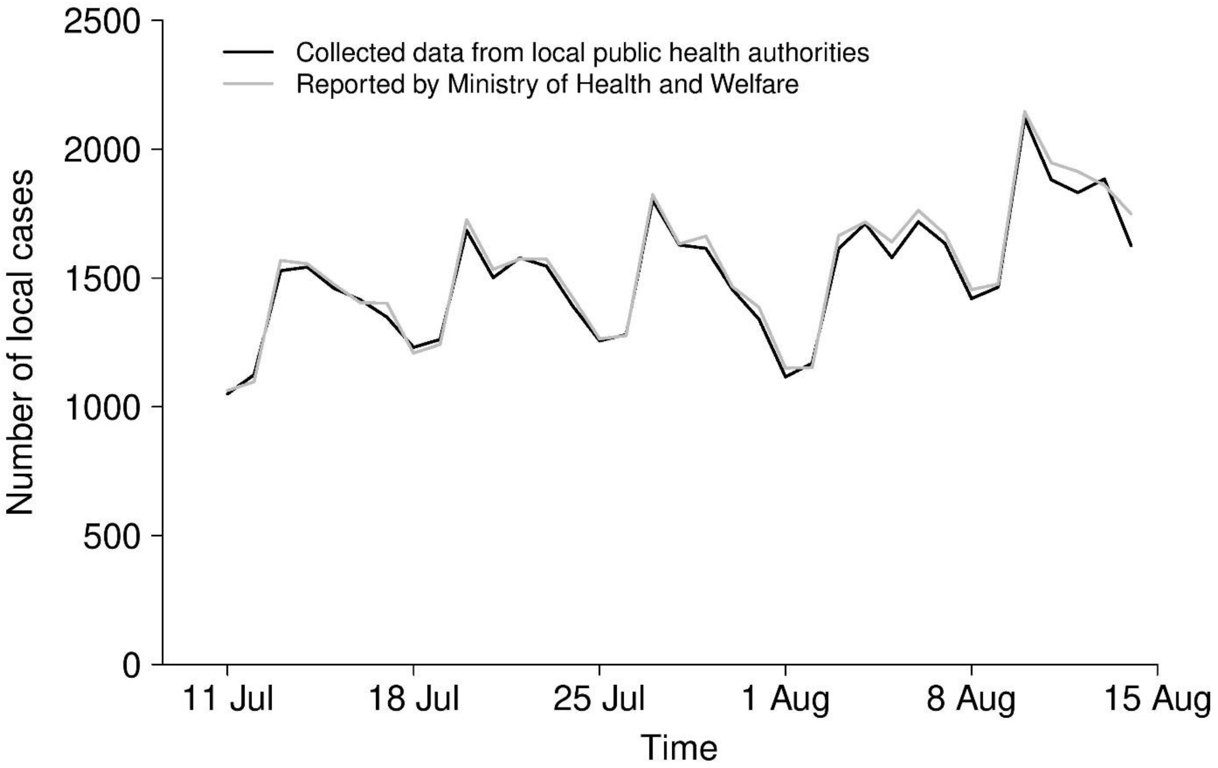


**Appendix Figure 2**. Daily number of COVID-19 cases from the collected data and reported number by the Korean Ministry of Health and Welfare in South Korea. The black line indicates the collected data from the local public health authority. The gray line indicates the daily reported number of COVID-19 cases from the South Korean central government.
